# How to handle the deceased body of COVID-19: an insight from Indonesian muslim burial handlers’ knowledge, perception, and practice

**DOI:** 10.1101/2020.08.03.20167593

**Authors:** Tri Bayu Purnama, Siti Khodijah, Idris Sadri

**Author notes:** Corresponding Authors: Tri Bayu Purnama, Faculty of Public Health, Universitas Islam Negeri sSumatera Utara Medan.

## Abstract

**Introduction:** Handling the remained body affected by COVID-19 in Indonesia is still an issue as it is done by community.

**Aim:** This study aims to explore the handler’s knowledge, perception, and practice in handling the remains.

**Method:** This study elaborates qualitative design in exploring the handler’s knowledge, perception, and practice in handling the remains dealing with universal precaution during the handling of the deceased body of COVID-19. The data is obtained through in-depth interview with guides.

**Result:** The majority of participants were acknowledged that the universal precaution is a precautionary conduct which remains general and nontechnical. Technicality on the universal precaution such as personal protective equipment and safety standards for infectious substance on the dead body are still incomprehensive. The perception of each handler of the body is still obscure. It was showed by cautious/apprehensive sense predominated the handlers in handling the dead body.

**Conclusion:** Improving the knowledge and strengthening the network of the handlers among the community play an important role in interrupting the chain of transmission of COVID-19 in the community.

## Introduction

COVID-19 pandemic remains persistent in Indonesia which is reported total number of COVID-19 93.657 cases and 4.576 mortal cases per July, 24th 2020. This pandemic spread over all provinces in Indonesia with the highest number of confirmed cases hit provinces entire Java Island. A plenty of medical measures associated with COVID-19 transmission have been conducted to interrupt new confirmed cases, new hospitalized cases and new death cases. Despite travel restriction, physical distancing, promoting health protocol and wearing mask, and school/office closure have been done, however, the number of COVID-19 cases is still rapidly increasing and driving to second wave in many provinces. Community attitude and behaviours due to rumours, fears and mistrust, social resistance might exacerbate the number of COVID-19 transmission case in Indonesia. It might deal with COVID-19 pandemic has transformed human behaviour drastically in very short term. A huge rumours, fear and denial of COVID-19 pandemic could be posed by burial rite of COVID-19 death body which is discarded from community in Indonesia.

Burial rites of COVID-19 cases is still a critical issues in many muslim communities (1). The burial procedure of the corpse during the COVID-19 pandemic disregards the procedures that have been long practiced by diverse religious and cultural communities (2). As a consequence, several cases of public rejection to the burial protocol of COVID-19 emerge in many parts of Indonesia for the previously mentioned reasons. The community respects the dignity of the burial rites as a final honor to the body. (3). Thus, there are many cases of forced withdrawal of COVID-19 cases by the deceased families which certainly threatens the safety of the human resources responsible for the burial process in the community. Under normal circumstances, the community bathes the body by being represented by the burial handlers and the family touches the body before it is buried (3). Furthermore, the transmission occurs in these activities and triggers a new cluster in transmission of the disease even though it is very unlikely that the transmission occurs (4).

Many studies found that there is asymptomatic COVID-19 cases circulating in the community and could transmited the disease to other with no symptom cases (5-8). This became worsen by the body of COVID-19 which was forcibly taken by a family which later is buried in the community (9). A huge gap of knowledge and varied and unsafe burial practices among muslim burial handlers could spread a new cluster of the disease and complicate to control andtrack the transmission. The objective of this study is to explore the difference knowledge and perception muslim burial handlers during the prolonged existence of the pandemic in Indonesia. We collected qualitative data through in-depth interview during the active COVID-19 transmission in North Sumatera Province, Indonesia. This study findings may include health message and strategy to prevent COVID-19 transmission in the community.

## Methods

### Study setting

A qualitative study was employed to explore knowledge, perception, and practice among muslim burial handlers on burial rites in COVID-19 cases. The exploration in the community and the potential for disease transmission within have been done by developing the phenomenon and perception of muslim burial handlers. Then, an investigation into the potential for developed knowledge and alternative solutions to interrupt the chain of transmission in the community or the potential of new cluster resources can be suggested to COVID-19 control policies.

This research instrument involved a research interview guide consisting of 4 parts. The first part (Part A) is the demographic characteristics of the informant such as sex, age, and length of active period as a corpse. The second part (Part B) is the knowledge of scavengers about universal vigilance in mortar scavenging activities. The third part (Part C) contains the perception of the body handlers about the vulnerability of the handlers’ body infected with infectious diseases. Part D or part four is a problem that might be done to increase public awareness of waste pickers and avoid infectious diseases in the perspective of waste pickers. A close investigation to this problem is managed by questioning the informant the unanswered questions. In addition, investigations were also carried out unfamiliar medical terms to the informant and replaced with local languages understood by the informant

### Participants

The target population in this study is the muslim burial handlers in North Sumatra. The limitation of this study are on the current situation and location of North Sumatra, this study interviewed 6 muslim burial handlers (participant) in accordance with the target population. All informants voluntarily agreed to participate in this study after obtaining an explanation of the study and agreed to the given consent form. The interviewers of this study are junior epidemiologists who have been trained to collect data with research instruments and are conducted with regard to health protocols established by the Indonesian Ministry of Health.

### Data Collection and Analysis

This research was conducted by interview and all participants’ answers were recorded by a sound recorder and verified in the interview matrix. The data is then analyzed by discussing content analysis by looking at the answers to open questions such as knowledge, reason, and perception which then propose a unique code for each answer of each informant. Thematic analysis is used to look at answer patterns and answer informants’ answers based on specific themes. The informants’ answers digged from closed questions and conclusions of open questions are compiled and transferred to percentages. The open-ended questions and the results of the investigation are compiled and presented in narrative form. This study was approved by the Faculty of Medicine, Islamic University of North Sumatra, Indonesia.

### Result

This study interviewed 6 muslim burial handlers with 5 out of 6 muslim burial handlers over 50 years old. More than half of the participants were male (4 out of 6) (table 1). The majority of participants (5/6) acknowledge universal precaution. The participants assume that universal precaution focuses on being caution at each stage of the burial process. However, technical universal precautions such as personal protective equipment and safety standards for biological agents can’t be clearly understood (0/6 participants).

> “universal precaution on this question was refered to general preparation during burial rites. (participant 1)

**Table 1.** Demographics of study participants

| Variables | N (%) |
| --- | --- |
| Sex |  |
| Male | 4 (66,7) |
| Female | 2 (33,3) |
| Age |  |
| < 50 yo | 1 (16,7) |
| ≥ 50 yo | 5 (83,3) |
| Had been trained |  |
| Yes | 0 (0,0) |
| No | 6 (100,0) |

Perception of COVID-19 among muslim burial handlers is very diverse. This can be seen from 3 out of 6 participants who answered the feeling of caution/fear burial rites. Participants answered the influence of television media was very outweighing affecting their point of view about COVID-19 transmission.

> “Initially, it was normal, just because more and more cases became more cautious and tend to be afraid of myself. If I get that close, maybe I will be paranoid, because I’m the type of person who is frightened, especially when my age is not young and there are comorbidities like heart disease and high blood pressure “(participant 1)

“A disease that can be transmitted to other people. Volunteering and praying for the best, support even though afraid “(participant 2)

> “I just feel normal, just a little worried because the media such as TV, moreover I live in the village so I think it’s safer. I will first ask the remains of the corpse what is the cause of death, and as much as possible use hygienic equipment so as to reduce transmission “(participant 5)

The muslim burial handlers suggests conducting community education about the stages of burial rites, health promotion and the availability of personal protective equipment to be important aspects in preventing transmission of COVID-19 (Figure 1). In the other group (3/6 informants) answered that they did not know what to do to prevent COVID-19 during the burial rites and make it fate.

> “In my opinion, knowledge about infectious diseases and their transmission as well as complete PPE from the prevention of these diseases” (participant 3).
>
> “Educate all citizens to be good at wahing body, so that he will wash his own family (participant 5)”.

**Figure 1.**
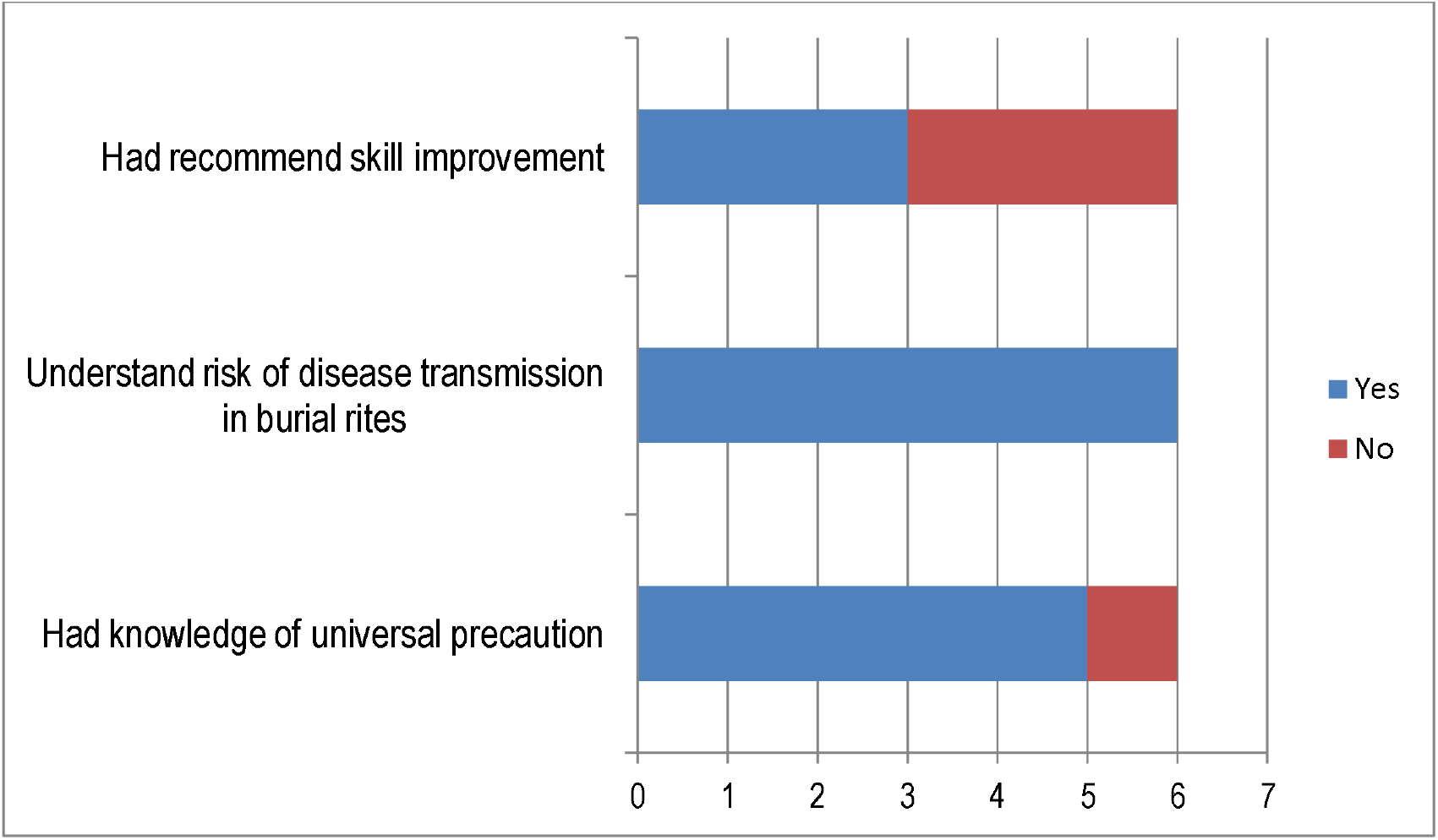
Thematic analysis of knowledge and perceive of burial participants.

## DISCUSSION

This research highlighted that muslim burial handlers is dominated by the elderly group. The elderly are a vulnerable group in transmission of COVID-19 and die with the presence of comorbidities from the COVID-19 (10). A study elsewhere found that *ghusl* (washing deceased whole body with soap and water) and *kafan* (shrouding of the deceased in white cotton sheets) as a part of burial rites were mostly conducted by elders in the muslim community due to the experience of the elderly group more expert and skillful compare with younger group (11). Some studies report that the elderly are vulnerable to COVID-19 due to immune responses and other comorbidities such as heart, respiratory and other terminal diseases (12). Age restriction as a muslim burial handlers remains important in reducing the risk of COVID-19 transmission. Lack of experience among younger muslim should be swiftly trainned with muslim community to conduct burial rites among muslim. Providing trainning through social media platforms could be invaluable strategy to organised teams of younger volunteers.

Burial: it is obligatory for Muslims to bury their dead as soon as possible after death, typically within 24 hours. It is recommended that the washed and shrouded body is buried without a coffin, with the face of the deceased facing Mecca.

The deceased muslim rites is obligatory for muslim to bury their dead within 24 hours which is started from ghusl (ritual washing deceased body), kafan (shrouding of the deceased), Salat al-Janazah (congregational prayer for the deceased), and burial as the end of deceased muslim rites (11). World health organization has recommended to conduct safe and dignified for dead body related to COVID-19 (2). In our study found that lack of information and personal protective equipment in the burial rites remains a crucial issue in this study during COVID-19 pandemic. The participants explained that there was no explanation about universal precautions for transmitting diseases from the corpse and self-protection equiptment in preventing infection from biological agents. The Ministry of Health and the Ministry of Religion of Indonesia issued regulations regarding the technical implementation of the burial rites (13). The availability of PPE is provided to muslim burial handlers in term of COVID-19 procedure in the hospital while there is no PPE support for muslim burial handlers in the community. Mistrust of burial rites procedure in the hospital forced community to refuse burial rites process in the hospital. Furthermore, community has requested muslim burial handlers in the community to process burial rites without any medical observation and fully personal protection equiptment. It would exacerbated the spectrum of COVID-19 pandemic geographically.

Muslim burial handlers did not get relevant information related to how to process burial rites in accordance with health and religious procedures. In addition there was a rejection due to misperception in the community related to the process of corpse by the COVID-19 procedure which was deemed incompatible with religious and cultural aspects before the pandemic occurred (14,15). The community tends to understand that last respects to humans is done by doing a good burial rites and being attended by a large number of participants (9). However, this action has an impact on the mode of transmission of infectious diseases. Research in Sierra Lione explains that improper burial rites and involving large numbers of people has an impact on the new cluster of Ebola cases (4). Promoting community to obey health protocols including health protocols in the past review of the burial rites becomes crucial in order to prevent the spread of the disease (16).

## Conclusion

The limited understanding and perception of muslim burial rites is a new issue in preventing the new COVID-19 cluster in the community. This is in line with the increasing reports of public refusal to process corpses with the COVID-19 procedure. Increased knowledge and strengthening of the scourge network in the community are important to break the chain of transmission of COVID-19 in the community.

## Data Availability

Data are available by requested

## Notes

### Competing Interest Statement

The authors have declared no competing interest.

### Funding Statement

No funding is available for this study

### Author Declarations

This study was approved by the Faculty of Medicine, Islamic University of North Sumatra, Indonesia

